# Animal husbandry and environmental conditions are associated with cefotaxime-resistant *Escherichia coli* in yard soil in peri-urban Malawi

**DOI:** 10.64898/2026.03.24.26349245

**Authors:** Emma Budden, Caitlin G Niven, Benjamin Clark, Emily Floess, Blessings Chirwa, Monica Matekenya, Stella Cadono, John Chavula, Victor Chisamanga, Aubrey Dzinkambani, Chisomo Kaponda, Neema Ngondo, Norah Patterson, Sheena Symon, Brighton Austin Chunga, Rochelle H Holm, Petros Chigwechokha, Francis L. de los Reyes, Cassandra L Workman, Angela Rose Harris, Ayse Ercumen

## Abstract

Soil is an important reservoir for antimicrobial resistance (AMR) and increasingly recognized as a pathogen transmission pathway. While studies have detected AMR in soil in various settings, dominant contributors to domestic soil contamination with antimicrobial-resistant organisms in low-income countries remain unidentified. We conducted a cross-sectional study with 237 households in southern Malawi, specifically peri-urban Bangwe near Blantyre, to identify factors associated with the abundance of cefotaxime*-*resistant *E. coli* in yard soil. Enumerators employed structured surveys and sampled 30 cm^2^ of yard soil per household. We used IDEXX Quanti-Tray/2000 with Colilert-18 and cefotaxime supplement to enumerate the most probable number (MPN) of cefotaxime-resistant *E. coli* per dry gram of soil. We conducted multivariable regression to assess associations between the abundance of cefotaxime-resistant *E. coli* and household sanitation, animal ownership and management, child health and antibiotic use, and weather. Of 228 soil samples, 68% harbored cefotaxime-resistant *E. coli* at a mean of 0.90 log_10_-MPN/dry gram. Compared to households without animals, households had approximately 0.50-log lower mean cefotaxime-resistant *E. coli* abundance in soil if animals were enclosed at night and 0.50-log higher abundance if they were not (p-values<0.005). Additionally, samples had approximately 0.70-log lower mean cefotaxime-resistant *E. coli* abundance if soil was dry at the time of collection and if it came from a household in the top wealth quintile (p-values<0.005). Daytime animal confinement, household sanitation, child health, antibiotic use, rainfall, temperature and ambient humidity were not associated with cefotaxime-resistant *E. coli* abundance. Findings suggest that animal husbandry and soil moisture had stronger associations with cefotaxime-resistant *E. coli* in soil compared to sanitation or antibiotic use. These results underscore the importance of a One Health approach and the relevance of domestic animals and environmental factors to AMR in soil. Studies should quantify soilborne AMR exposure and evaluate associations with specific animal management/enclosure practices.

## Introduction

Antimicrobial resistance (AMR) is a leading threat to public health globally. In 2019, AMR was associated with 1.27 million deaths [1]. By 2050, it is estimated that this will reach 10 million deaths per year [2], while others predict 2 million deaths directly caused by AMR and 8 million deaths related to AMR [3]. Infections caused by antibiotic resistant organisms (AROs) are increasingly difficult to treat with available antibiotics and are associated with increased risk of death and complications, longer duration of illness, and increased healthcare costs [4–7]. Community colonization of AMR is particularly high in low-income countries. In sub-Saharan Africa, intestinal colonization with extended-spectrum beta-lactamase (ESBL) producing *E. coli* ranged from 5 to 59% of the population, where children <5 years in the Central African Republic had the highest colonization at 59% [8]. Further, a study conducted in 2023 found that 89% of infants in a Kenya hospital were colonized with ESBL-producing bacteria [9]. In many low-income countries, regulations on antibiotic use are limited [8], and clinically-relevant antibiotics are often available without restriction, both for human use and animal husbandry [10]. Frequent antibiotic use, including self-prescribing or over-prescribing by healthcare professionals [11] and therapeutic or sub-therapeutic use for animals [12] is a known driver of AMR.

Poor water and sanitation infrastructure, and contaminated environments further facilitate the spread of AROs and antimicrobial resistance genes (ARGs) between reservoirs and hosts via horizontal gene transfer [13–15]. Antibiotic residues, AROs, and ARGs can be shed in human feces, leading to environmental dissemination of AMR in settings with open defecation or unsafe sanitation infrastructure [16,17]. Similarly, when domestic animals defecate near the dwelling, antibiotic residues, AROs and ARGs from animal fecal waste can be disseminated into soil and other environmental compartments and further proliferate via horizonal gene transfer [18,19]. Environmental compartments are recognized as reservoirs for AMR where mobile genetic elements can be exchanged between native communities and pathogens deposited in the environment via human or animal fecal waste [20,21]. Environmental conditions, such as temperature and precipitation patterns, may affect the proliferation of ARGs [22]. Additionally, while ARGs can spread naturally in the environment, anthropogenic disturbances can accelerate their dissemination by adding selective pressure [16,23]. For example, ARG exchange between organisms, such as through horizontal gene transfer, can be triggered by co-selectors, such as heavy metals, pesticides, and microplastics [23].

As one of the most diverse matrices for microbial activity, soil is a particularly important environmental reservoir for ARG proliferation [24] because it can offer an efficient environment for ARGs to transfer between soil-inhabited bacteria and clinically-relevant pathogenic organisms, such as *E. coli*, *Klebsiella pneumoniae*, and *Salmonella enterica* [25,26] (Figure 1). In rural Bangladesh, a study detected cefotaxime-resistant *E. coli* on 89% of soil floors [27]. Resistance against at least one antibiotic was detected in *E. coli* isolates from yard soil samples in 42% of isolates in Bangladesh [28] and 33% of isolates in Peru [29]. Further, a recent study in Nigeria found ESBL-producing *E. coli* in 68% of farm soil samples [30], and a study conducted along a river basin in Tanzania found that 14% of samples positive for *E. coli* harbored ESBL-producing organisms [31].

**Figure 1.**
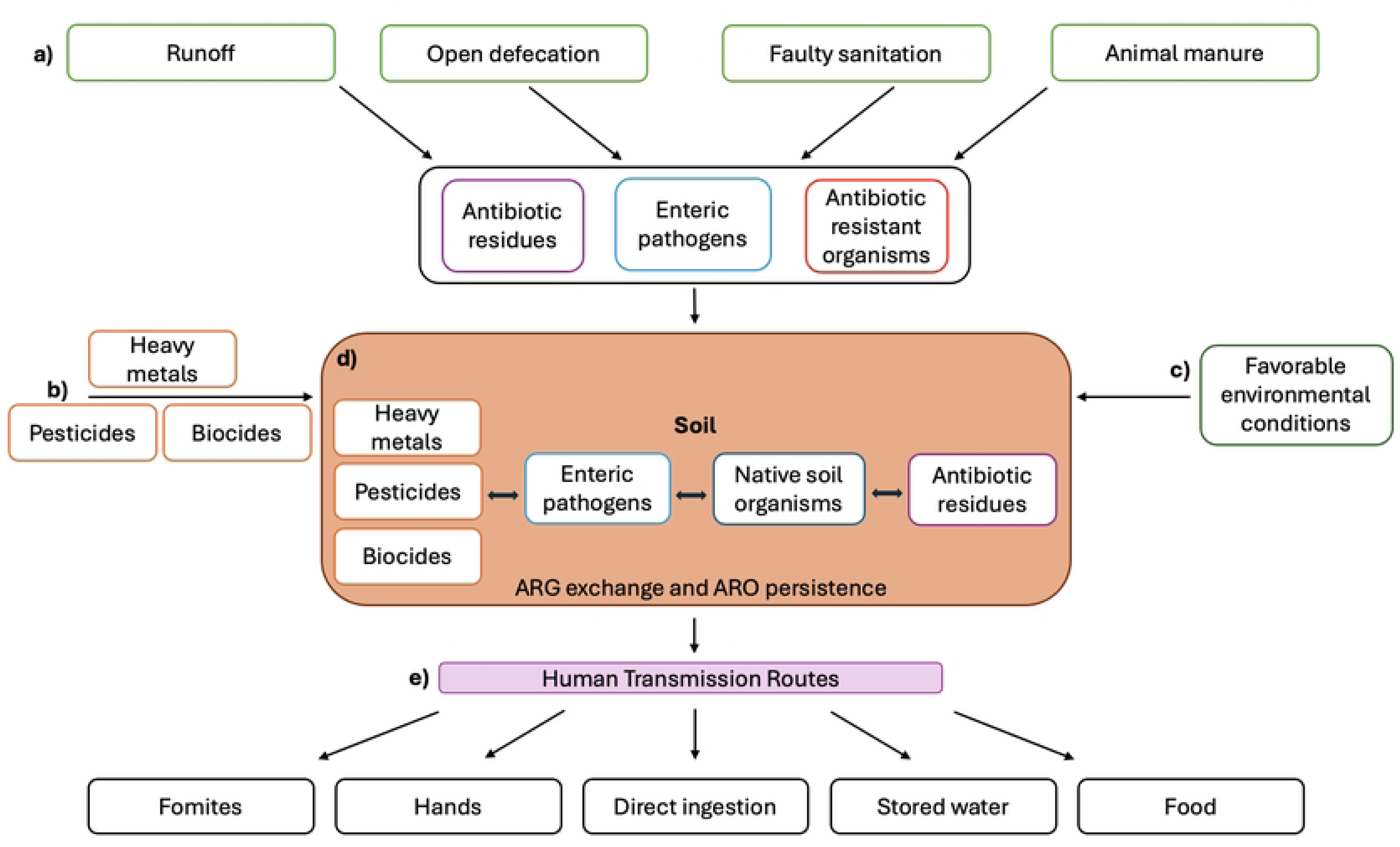
a) Fecal waste streams, including runoff, open defecation, faulty sanitation, and unsafely managed animal manure, can deliver enteric pathogens, antibiotic residues and antimicrobial-resistant organisms (AROs) to soil; b) human-induced disturbances may result in co-selectors, such as heavy metals, pesticides, and biocides in soil; c) environmental variables, such as moisture and elevated temperatures, can support favorable conditions for antibiotic resistance gene (ARG) exchange and ARO persistence; d) native soil organisms interact with delivered pathogens, antibiotic residues, AROs, heavy metals, and chemicals, and a confluence of biological and co-selectors can trigger ARG exchange and support ARO persistence in soil; e) human transmission pathways of AROs from soil include direct ingestion of soil and/or indirect ingestion from soil-contaminated hands, fomites, stored water and/or food.

Soil contaminated with fecal waste can be directly ingested by young children via mouthing behavior or indirectly via contaminated food, water, fomites, and/or hands, which can transfer fecal pathogens to hosts [16,32,33] (Figure 1). Direct soil consumption among children has been correlated with increased diarrheal diseases [34,35]. Early childhood behaviors, such as mouthing soiled hands or objects, further elevates the risk of young children contracting enteric diseases through indirect soilborne routes [36]. In a Monte Carlo simulation utilizing data from rural Bangladesh, 40% of children between six months and two years of age were found to ingest soil [37]. While no studies have assessed whether soil exposure is associated with increased AMR colonization, it is plausible that AROs and ARGs can be transmitted to hosts via soilborne pathways. For example, in a 2022 study in Malawi, having a toilet with a soil floor was associated with increased odds of having ESBL-producing *E. coli* in stool samples collected from household members [38].

Sanitation practices have been explored as a determinant of both overall fecal contamination and AMR contamination in domestic soils, but the evidence is mixed for fecal indicators and scarce for AMR. A study in Tanzania found that having an improved versus unimproved pit latrine was not associated with fecal indicator bacteria in yard soil; soil at the entrance to the home had some of the highest concentrations of fecal indicator bacteria of all sampled locations, including the latrine entrance [39]. It was hypothesized that latrine superstructure may influence this finding because most latrines had open roofs, which may result in bacterial inactivation by sunlight [39]. Similarly, a 2018 study in rural Bangladesh found that having an improved versus unimproved pit latrine was not associated with *E. coli* or ESBL-producing *E. coli* in yard soil [28]. A randomized sanitation intervention providing improved latrines in rural Bangladesh did not reduce *E. coli* levels in yard soil; though this study did not evaluate resistance [40]. Sanitation improvements are recommended in AMR control programs [41], and a better understanding of how sanitation infrastructure/practices influence AMR in soil can inform AMR mitigation strategies.

Animal husbandry is a major source of fecal contamination in the environment and plays an important role in disseminating AMR [13]. Studies in China and Italy found that hog feces and soil from hog farms have high rates of detection for ARGs, and there is similar evidence for poultry and cattle farms in studies from China, pinpointing animal feces as a source of AMR in animal production facilities [42–46]. Other studies have investigated fecal indicator bacteria and AMR in informal backyard animal husbandry settings, a common practice in low- and middle-income countries where animals are raised within the premises. In Bangladesh, the presence of chickens, cows, goats, and sheep in the household compound increased *E. coli* abundance in yard soil, and chicken ownership in particular was correlated with significantly higher abundance [32]. Also in Bangladesh, cefotaxime-resistant *E. coli* on indoor soil floors had a strong correlation with owning domestic animals and unsafe animal management practices, such as letting animals roam free in the home and keeping them inside the home at night [27]. A study found that, in low- and middle-income countries, animal roaming in or near living spaces allows for the exchange of ARGs through environmental compartments, and soil in particular plays a critical role as a mediator in AMR transmission between humans and domestic animals [47]. Given that backyard animal husbandry is a common form of livelihood in low-income countries, the influence of specific animal management practices (e.g. roaming, daytime and nighttime confinement) on the environmental dissemination of AMR needs further study.

Prior research has documented that microbial growth is influenced by environmental and weather factors. Higher soil moisture content heavily influences the proliferation of *E. coli* and ESBL-producing *E. coli* in soil [28]. A 2024 study found a higher prevalence of beta-lactamase genes in soils where manure had been applied under higher moisture conditions [48]. There is evidence that temperature and sunlight exposure also influences *E. coli* concentration in soil [39,49]. In a study in rural Tanzania, lower concentrations of fecal indicator bacteria were found in soil that was in direct sunlight compared to in shade or partial sunlight [39]. Additionally, *E. coli* inoculated into soils from Lake Superior (bordered by the United States and Canada) and observed under varying temperature conditions were found to grow more successfully in temperatures ≥30 °C, with the most growth occurring at 37 °C [49]. Further evidence is needed on the influence of environmental conditions (e.g. sunlight, rainfall, ambient temperature, ambient humidity, soil moisture) on antimicrobial-resistant organisms.

In this study, we aimed to quantify cefotaxime-resistant *E. coli* in yard soil among peri-urban Malawian households. Cefotaxime is a broad-spectrum cephalosporin antibiotic that is commonly used in clinical settings [50] and a known indicator for ESBL-producing bacteria, which are of particular concern due to their ability to facilitate multi-drug resistance against clinically relevant antimicrobials [50]. Specifically, ESBL-producing *E. coli* forms the basis of WHO’s global AMR surveillance in environmental compartments [51]. Here, we used a One Health approach to assess associations between the abundance of cefotaxime-resistant *E. coli* in yard soil and a range of human, animal and environmental factors, including sanitation, animal management practices, antibiotic use by humans and animals, and weather-related variables.

## Methods

### Study design

From June 20, 2024 through July 14, 2024, we conducted a cross-sectional study with 237 households in Bangwe, a peri-urban municipality in Blantyre, Malawi. We aimed to enroll households in geographical clusters, corresponding to informal neighborhood delineations [52]. Starting at random points within each targeted area, enumerators travelled in each cardinal direction to screen every third household for eligibility. If the household had a child <5 years old and a caretaker at least 18 years of age consenting to participate, the household was enrolled in the study via written consent. In all enrolled households, enumerators administered a structured survey and collected samples of yard soil.

### Ethical considerations

We fulfilled all Institutional Review Board requirements through Mzuzu University Institutional Review Ethics Committee (#MZUNIREC/DOR/24/38) and the University of North Carolina at Greensboro (UNC-Greensboro IRB #: 20-0245). A caretaker over the age of 18 provided written informed consent to participate. Permission to conduct the study was approved by Blantyre City officials as well as local leaders in Bangwe township.

### Data collection

Enumerators from Mzuzu University and the Malawi University of Science and Technology were trained to administer a structured questionnaire in Chichewa using Open Data Kit (ODK). The questionnaire included questions and spot check observations of household water and sanitation indicators drawn from the Joint Monitoring Programme’s (JMP) Core Questions on Drinking Water, Sanitation and Hygiene for Household Surveys (e.g. water source, latrine type and access, child defecation) [53], animal ownership and management (e.g. types of domestic animals, daytime and nighttime confinement), caregiver-reported child health (e.g. enteric/respiratory infection symptoms, antibiotic use), environmental factors (e.g. sampled soil in sunlight, soil wetness) and socio-demographic indicators (e.g. assets, expenditures, education, household size, floor material). The questionnaire also recorded caregiver-reported child soil exposure behaviors. Ambient temperature and humidity were measured using five EXTECH Instruments Humidity/Temperature Dataloggers RHT10 placed in various locations around Bangwe throughout the data collection period, recording ambient temperature and relative humidity every ten minutes. Additionally, we collected publicly available temperature and humidity data from Weather API, which recorded ambient temperature and humidity every three hours in Blantyre [54], the closest city to the study area.

### Soil sample collection

Enumerators trained in aseptic technique identified a courtyard area immediately adjacent to the entrance of the household, avoiding stepping into the sampling area. Two stainless steel corner stencils and a stainless-steel spoon sterilized with 10% bleach and 70% ethanol were placed on the soil to measure a 30 x 30 cm area. The spoon was used to scrape the topsoil 10 times vertically and 10 times horizontally, and then to transfer the collected soil into a sterile Whirlpak bag. The Whirlpak bag was placed in a secondary plastic bag to avoid cross-contamination, stored in a cooler with ice, and transported to the Biological Sciences Laboratory at the Malawi University of Science and Technology. A field blank was administered every other day. At the first household enumerators visited, enumerators opened a Whirlpak bag containing sterile DI water, stirred the sanitized sampling spoon ten times, closed the bag, and returned it to the lab with the soil samples.

### Enumeration of cefotaxime-resistant E. coli

Samples were processed at the Malawi University of Science and Technology within five hours of collection. The Whirlpak bag containing the soil sample was shaken to homogenize. Then, 4 g of soil and 40 mL of sterile DI water were added to a 50 mL falcon tube and vortexed for 2 minutes. 10 mL of the homogenized sample was retrieved without letting the soil settle and added to a sterile 100 mL Whirlpak bag containing 90 mL of sterile DI water, creating a 100 mL aliquot that contained 1 g of soil. 80 µL of 5 mg/mL filter-sterilized cefotaxime was added to the 100 mL solution, for a final cefotaxime concentration of 0.004 mg/mL [55], followed by adding Colilert-18 media. Field blanks and 10% lab blanks were analyzed using the same procedure, using 100 mL of field blank sample and 100 mL of sterile DI water, respectively. Two aliquots were analyzed for each blank, one with and one without cefotaxime. All Whirlpak bags were gently inverted to mix and, once the media dissolved, the contents were added into a Quanti-Tray/2000 and sealed. Sealed trays were incubated at 35 °C for 20-22 hours and visually enumerated for the most probable number (MPN) of cefotaxime-resistant *E. coli* using a UV lamp. Each soil sample was also assessed for its moisture content. An approximately 5 g aliquot of soil was first weighed to record the wet mass [32]. Samples were then placed into a drying oven at 110 °C for 24 hours. After 24 hours, samples were removed from the oven, and the dry mass was recorded. The moisture content was used to calculate the MPN of cefotaxime-resistant *E. coli* per dry gram of soil.

### Statistical analysis

We tabulated household-level descriptive statistics on socio-demographics, water, sanitation, and hygiene conditions, animal ownership and management practices, child health and antibiotic use, and soil exposure. We generated a binary variable for the prevalence of cefotaxime-resistant *E. coli* and log_10_-transformed the MPN values per dry gram. Non-detect samples were assigned 0.5 MPN/wet gram of soil (half the lower detection limit) [56], and samples exceeding the upper limit of quantification of 2419 MPN were assigned 2420 MPN/wet gram prior to calculating the MPN/dry gram and taking the logarithm. We tabulated the mean log_10_-transformed MPN of cefotaxime-resistant *E. coli* in yard soil across cross-categories of animal ownership and husbandry practices and cross-categories of latrine type and user load. We used bivariate and multivariable regression to assess associations between household and environmental variables and the log_10_-transformed MPN and prevalence of cefotaxime-resistant *E. coli*. We considered the following independent variables in bivariate models.

#### Sanitation practices

We generated binary variables for whether: any feces were observed within 2×2 meters of the soil sampling area; the household had an improved latrine according to the JMP definition [57]; the household had a flush/pour flush latrine; the latrine was utilized by a single household or shared by multiple households; and children in the household open defecated.

#### Animal ownership and management

We generated binary variables for whether: the household owned animals; animals were kept in the compound (any animals combined as well as specific animal types); and any animal feces were observed in the compound. Among households that owned animals, we generated binary variables for whether animals were enclosed during the day and at night, and whether the household reported giving antibiotics to animals owned (any animals combined as well as specific animal types). Animals were considered enclosed if they were kept in a basket or cage, animal-only enclosure that they could not leave, or an animal-only enclosure that animals could leave freely. Additionally, we generated a categorical variable for animal cohabitation intensity: household owns animals that are not enclosed, household owns animals that are enclosed, household does not own animals.

#### Environmental factors

We generated binary variables for whether the soil sampling area was in sunlight and whether it was wet at the time of collection as observed by the enumerator. We performed a two-tailed t-test to determine if the enumerator-recorded observation of soil wetness correlated with the soil sample’s measured moisture content. We tabulated the daily average temperature and humidity using data from the loggers. For study dates where only a subset of loggers was online for 24 hours because of installation/de-installation partway through the day (June 25), we only used data from the loggers that had data for the full 24-hour period. For study dates with no available logger data (June 20, 21, 24), we calculated the daily average temperature and humidity using data from Weather API (weatherapi.com/history/q/blantyre-1598090). We created tertiles of daily average temperature and humidity across the study period and generated binary variables for the top vs. bottom two tertiles.

#### Child illness and antibiotic use

We generated binary variables for whether any child <5 years in the household had symptoms of caregiver-reported diarrhea, acute respiratory infection (ARI), ARI with fever, or fever in the last seven days, and if any child <5 years in the household used antibiotics in the last four weeks. Diarrhea was defined as 3 or more loose stools in a 24-hour period. ARI was defined as respiratory symptoms, such as persistent coughing accompanied with shortness of breath, panting, or wheezing [58].

Bivariate models were run for each independent variable. Variables with a p-value of <0.20 in bivariate models were shortlisted for inclusion in a multivariable regression model. We estimated variance inflation factors to assess collinearity between the four weather-related variables (sampling area in sunlight, sampling area wet, average ambient temperature, average ambient humidity) before including these variables in multivariable models. The multivariable models also controlled for household’s primary water source, highest education level reached by any household member, household size, weekly household expenditure, indoor floor material and asset-based household wealth quintile. Household’s primary water source was categorized according to the JMP definition of improved vs. unimproved water sources [57].The indoor floor material was categorized as improved (cement/concrete, tile) vs. unimproved (earth/mud). The asset-based wealth quintile was calculated using a principle component analysis based on household-reported items owned [59]. These additional variables were selected for inclusion in multivariable models as potential confounders because household-level water service and socio-demographic factors are expected to influence AMR occurrence in the domestic environment [60,61]. Models only included confounders with sufficient variation in the sample (greater than 5% prevalence in each stratum).

We used generalized linear models to estimate associations. Models with continuous outcomes (log_10_-MPN of cefotaxime-resistant *E. coli*) used a Gaussian error distribution and an identity link to estimate the difference in log_10_-transformed MPN between groups of households, and models with binary outcomes (prevalence of cefotaxime-resistant *E. coli*) used a Poisson distribution and a log link to estimate prevalence ratios. Models accounted for geographic clustering of enrolled households using robust standard errors. All analyses were conducted in R version 4.3.3 through Rstudio version 2024.12.0+467.

## Results

### Participant characteristics

Between June and July 2024, 237 households were enrolled in the study, which were grouped into 38 independent spatial clusters based on their GPS coordinates [52]. Respondents were an average age of 31.1 years old, and households included an average of 1.2 children <5 years, 1.4 children aged 5-16 years, and 2.7 adults >16 years (S1 Table). 87% (205) of respondents reported being able to read and write, and the most common education level was secondary (partial or complete), both among respondents (50%, n=118) and all household members (63%, n=149) (S1 Table). Household average weekly household expenditure was 17.0 USD (S1 Table). Additionally, 69% (164) of households had electricity in the previous 7 days, 24% (56) owned a refrigerator, 70% (166) owned a mosquito net, and 87% (205) owned a mobile phone (S1 Table).

#### Water and sanitation practices

84% (200) of households had an improved primary water source, and the most common source was piped water outside the compound (29%, n=69) (S1 Table). Most households had a latrine, while 19% (46) of households had an improved latrine, and the most common latrine type was a pit latrine with slab (65%, n=154) (S1 Table). 71% (168) of respondents reported sharing their latrine with other households, with an average of 11 people per shared latrine (S1 Table). The primary place of defecation for children <5 years was in latrines (40%, n=95) and nappy/diapers (39%, n=92) (S1 Table). However, children <5 years were also reported to openly defecate in the compound in 25% (59) of households and outside of the compound in 5% (11) of households (S1 Table). The majority (65%, n=153) of respondents reported putting/rinsing child feces into the latrine (S1 Table). In 8% (20) of households, fecal waste was observed within 2×2 meters of the soil sampling area in the yard (S1 Table).

#### Animal ownership and management

Animals were owned by 24% (57) of households; the most common animal was chickens/poultry (16%, n=37) while other animals included dogs (9%, n=22), cats (5%, n=12) and sheep/goats (0.4%, n=1) (Table 1). Almost three quarters (75%, n=177) of households reported that animals lived in their compound, where a compound was defined as the land area where the household lives, including extended family members. The most common animal living in the compounds was chickens/poultry (61%, n=144) while dogs, cats, sheep/goats, and pigs were also reported (Table 1). Households owned an average of 1.8 animals, and an average of 7.1 animals lived in the compound (Table 1). Animals were observed within 2×2 meters of the soil sampling area in the yard in 7% (16) of households, and animal feces were observed in the compound in 37% (21) of households (Table 1).

**Table 1.** Animal ownership and management practices among enrolled households.

| Animal ownership | N=237 |
| --- | --- |
| Household keeps animals/livestock, % (n) |  |
| Any animal | 24.1 (57) |
| Chickens/poultry | 15.6 (37) |
| Sheep/goats | 0.4 (1) |
| Dogs | 9.3 (22) |
| Cats | 5.1 (12) |
| Other | 0.8 (2) |
| Animals/livestock live in compound, % (n) |  |
| Any animal | 74.7 (177) |
| Chickens/poultry | 60.8 (144) |
| Sheep/goats | 2.1 (5) |
| Pigs | 0.4 (1) |
| Dogs | 46.0 (109) |
| Cats | 8.0 (19) |
| Number of animals kept by household, mean (SD) |  |
| Any animal | 1.8 (6.3) |
| Chickens/poultry | 1.5 (6.0) |
| Sheep/goats | 0 (0.1) |
| Dogs | 0.2 (0.7) |
| Cats | 0.1 (0.4) |
| Other | 0 (0.1) |
| Number of animals living in compound, mean (SD) |  |
| Any animal | 7.1 (8.7) |
| Chickens/poultry | 5.0 (6.9) |
| Sheep/goats | 0.1 (0.8) |
| Pigs | 0.2 (3.2) |
| Dogs | 1.7 (2.7) |
| Cats | 0.1 (0.6) |
| Animals observed within 2x2 meters of soil sampling area, % (n) | 6.8 (16) |
| Animal feces observed in compound, % (n) | 36.8 (21) |
| Animal management practices % (n)<sup>a</sup> | N=57<sup>b</sup> |
| Animals kept during the day, % (n)<sup>a</sup> |  |
| Free roaming outdoors | 49.1 (28) |
| Free roaming indoors | 47.4 (27) |
| All enclosed | 12.3 (7) |
| Other | 1.8 (1) |
| Animal ownership among households that enclose animals during the day, % (n) | N=7<sup>c</sup> |
| Own only chickens/poultry | 57.1 (4) |
| Own chickens/poultry and other animals | 28.6 (2) |
| Own only other animals (sheep/goats, dogs, cats) | 14.3 (1) |
| Animals kept at night, % (n)<sup>a</sup> |  |
| Free roaming outdoors | 26.3 (15) |
| Free roaming indoors | 66.7 (38) |
| All enclosed | 21.1 (12) |
| Other | 1.8 (1) |
| Animal ownership among households that enclose animals at night, % (n) | N=12<sup>d</sup> |
| Own only chickens/poultry | 50.0 (6) |
| Own chickens/poultry and other animals | 41.7 (5) |
| Own only other animals (sheep/goats, dogs, cats) | 8.3 (1) |
| Household used antibiotics for animals in last 4 weeks, % (n) | N=57<sup>b</sup> |
| Any animal | 28.1 (16) |
| Chickens/poultry | 14.0 (8) |
| Dogs | 12.3 (7) |
| Cats | 1.8 (1) |

<sup>a</sup> Respondent could select multiple answers for these questions.
<sup>b</sup> Asked among 57 households that keep animals.
<sup>c</sup> Asked among 7 households that enclose animals during the day.
<sup>d</sup> Asked among 12 households that enclose animals at night.

Among the 57 households that owned animals, during the day animals roamed free outdoors in 49% (28) of households, roamed free indoors in 47% (27) of households, and were enclosed in 12% (7) of households (Table 1). At night, animals roamed free outdoors in 26% (15) of households, roamed free indoors in 67% (38) of households, and were enclosed in 21% (12) of households (Table 1). Among households that used a daytime enclosure, 57% (4) had only chickens/poultry, 29% (2) had chickens/poultry and other animals, and 14% (1) had only other animals, while among households that used a nighttime enclosure, 50% (6) had only chickens/poultry, 42% (5) had chickens/poultry and other animals, and 8% (1) had only other animals (Table 1). In the last four weeks, 28% (16) of households gave antibiotics to any animal in the household, most commonly to chickens/poultry (Table 1).

#### Child illness, antibiotic use and soil exposure

Among 292 children <5 years in enrolled households, caregivers reported that 16% (48) of children had diarrhea, 46% (134) had an ARI, 24% (70) had an ARI with fever, and 33% (97) had a fever in the last 7 days. In the last four weeks, 33% (97) of children used antibiotics; among those children, the average frequency of use was 1.2 antibiotic courses with an average of 1.8 doses taken (S2 Table). The most common antibiotics were amoxicillin (part of penicillin class) and trimethoprim/sulfamethoxazole (part of sulfonamide class) (S2 Table). Antibiotic recommendations most commonly came from doctors (54%, n=52), followed by self-prescription (29%, n=28) and pharmacists (17%, n=16) (S2 Table). The most common reason why the child stopped taking antibiotics was that the child felt better (47%, n=46) (S2 Table). Among the children who took antibiotics in the last three months, caregivers reported that the antibiotic was ineffective for 25% (24) of them, and of these 24 children, 71% (n=17) took an additional antibiotic to improve (S2 Table). Additionally, among children <5 years, 90% (263) played, 74% (217) ate, 49% (143) slept, and 25% (74) crawled on the ground outside; 9% (27) of children spent no time on the ground outside (S2 Table).

### Cefotaxime-resistant E. coli in yard soil samples

Yard soil samples were collected from 228 of 237 enrolled households; 9 households declined sample collection. 68% (156) of samples harbored cefotaxime-resistant *E. coli* at a mean log_10_-MPN of 0.90 (standard deviation [SD]=1.1), corresponding to a geometric mean of 8.0 MPN/dry gram (Table 2).

**Table 2.**
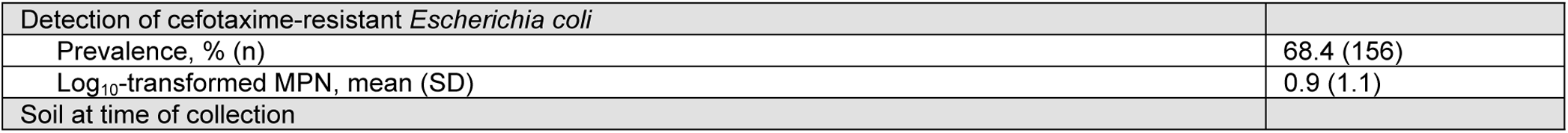
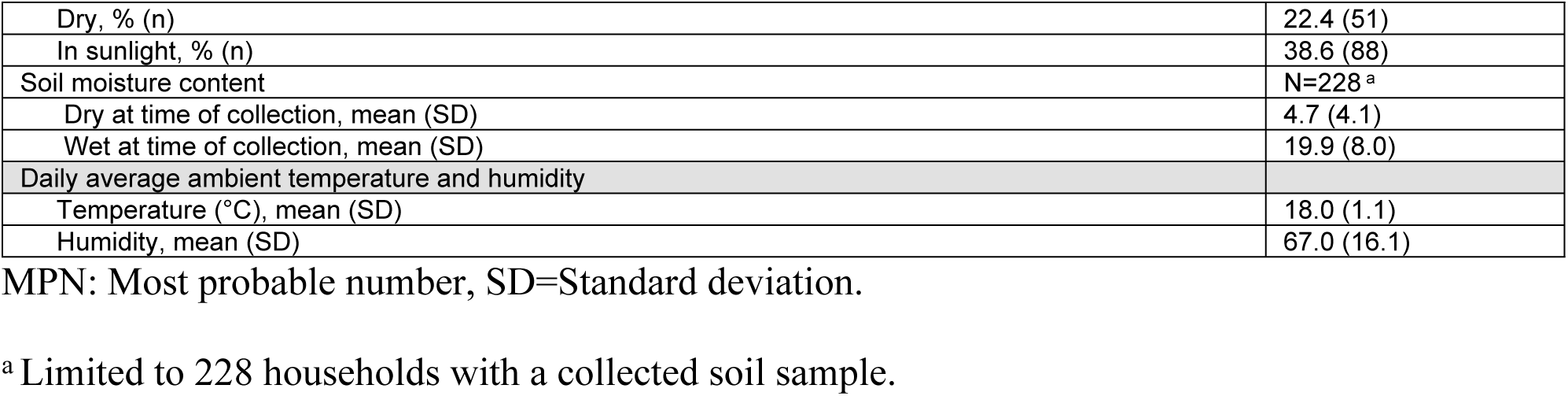
Soil and environmental characteristics.

| Detection of cefotaxime-resistant *Escherichia coli* |  |
| --- | --- |
| Prevalence, % (n) | 68.4 (156) |
| Log<sub>10</sub>-transformed MPN, mean (SD) | 0.9 (1.1) |
| Soil at time of collection |  |
| Dry, % (n) | 22.4 (51) |
| In sunlight, % (n) | 38.6 (88) |
| Soil moisture content | N=228<sup>a</sup> |
| Dry at time of collection, mean (SD) | 4.7 (4.1) |
| Wet at time of collection, mean (SD) | 19.9 (8.0) |
| Daily average ambient temperature and humidity |  |
| Temperature (°C), mean (SD) | 18.0 (1.1) |
| Humidity, mean (SD) | 67.0 (16.1) |

MPN: Most probable number, SD=Standard deviation.
<sup>a</sup> Limited to 228 households with a collected soil sample.

### Soil and weather conditions

At the time of collection, 39% (88) of soil samples were in sunlight and 22% (51) were dry (Table 2). When the sample was visually wet as reported by enumerators, the measured soil moisture content was significantly higher (20% for wet soil vs. 5% for dry soil, t-test p-value<0.05) (S1 Figure). The daily average temperature was 18.0°C, and the daily average relative humidity was 67% over the study period (Table 2).

### Household and environmental factors associated with cefotaxime-resistant E. coli in yard soil

Among cross-categories of animal type and animal enclosure practices, the abundance of cefotaxime-resistant *E. coli* in yard soil was highest among households that owned poultry and did not enclose their animals at night (mean log_10_-MPN: 1.49) and generally appeared higher when animals were not enclosed at night, regardless of the types of animals owned (S2 Figure). Among cross-categories of number of latrine users and latrine type, cefotaxime-resistant *E. coli* abundance in soil was high among households whose latrine was unimproved and used by an above-median (median=9) number of people (mean log_10_-MPN: 1.06), as well as among households whose latrine was not flush/pour flush and used by an above-median number of people (mean log_10_-MPN: 1.08). Cefotaxime-resistant *E. coli* abundance generally appeared higher among households where the latrine was used by multiple households. Unless the latrine was used by a single household, households with an unimproved latrine appeared to have higher cefotaxime-resistant *E. coli* levels compared to households with an improved latrine.

In bivariate regression, households had a statistically significant lower abundance of cefotaxime-resistant *E. coli* per dry gram of yard soil if owned animals were reported to be enclosed at night (Δlog_10_: -1.15 (-1.64, -0.67); p-value<0.0005), if the family’s latrine was used by a single household rather than shared with other households (Δlog_10_: -0.43 (-0.74, -0.13); p-value=0.01), if the soil was dry at time of collection (Δlog_10_: -0.89 (-1.24, -0.54); p-value<0.0005), or if the ambient temperature was in the top tertile (Δlog_10_: -0.29 (-0.56, -0.01); p-value=0.05) (Table 3). While not statistically significant, households also appeared to have lower abundance of cefotaxime-resistant *E. coli* if they had an improved latrine, any child had in the household had a fever in the last week or used antibiotics in the last four weeks (Table 3). On the other hand, households appeared to have a higher abundance of cefotaxime-resistant *E. coli* per gram of yard soil if the household owned any animal or owned chickens/poultry (Table 3). The following variables were not associated with cefotaxime-resistant *E. coli* abundance in yard soil: utilizing a flush/pour flush latrine, children in household openly defecating, feces or animals observed near sampling area, owning dogs/cats, compound keeping animals, animals enclosed during the day, household giving antibiotics to any animal, ambient humidity, and children having diarrhea, ARI, and ARI with fever (Table 3).

**Table 3.** Bivariate associations between household environmental characteristics and abundance of cefotaxime-resistant *E. coli* in yard soil. Bolded values indicate associations with p-value <0.20.

|  | Yes |  | No |  |  |  |
| --- | --- | --- | --- | --- | --- | --- |
|  | N | log<sub>10</sub>-MPN<br>Mean (SD) | N | log<sub>10</sub>-MPN<br>Mean (SD) | Dlog<sub>10</sub>-MPN<br>(95% CI) | p-value |
| **Sanitation** |  |  |  |  |  |  |
| Improved latrine | 45 | 0.60 (1.09) | 183 | 0.98 (1.11) | **-0.38 [-0.82, 0.06]** | **0.09** |
| Flush/pour flush latrine | 20 | 0.66 (1.40) | 208 | 0.93 (1.09) | -0.27 [-0.83, 0.30] | 0.35 |
| Latrine used by single household | 65 | 0.60 (1.05) | 161 | 1.04 (1.12) | **-0.43 [-0.74, -0.13]** | **0.01** |
| Children in household openly defecate | 67 | 1.04 (1.19) | 161 | 0.85 (1.08) | 0.20 [-0.19, 0.59] | 0.32 |
| Feces observed within 2x2 m of soil sampling area | 18 | 0.81 (1.08) | 210 | 0.91 (1.12) | -0.10 [-0.68, 0.48] | 0.73 |
| **Animal ownership and management** |  |  |  |  |  |  |
| Household owns |  |  |  |  |  |  |
| Animals | 55 | 1.07 (1.14) | 173 | 0.85 (1.11) | **0.22 [-0.08, 0.51]** | **0.15** |
| Poultry | 35 | 1.07 (1.07) | 193 | 0.87 (1.12) | **0.20 [-0.04, 0.44]** | **0.10** |
| Dogs/cats | 30 | 0.85 (1.18) | 198 | 0.91 (1.11) | -0.06 [-0.51, 0.38] | 0.78 |
| Compound keeps |  |  |  |  |  |  |
| Animals | 172 | 0.92 (1.12) | 56 | 0.86 (1.11) | 0.06 [-0.29, 0.40] | 0.75 |
| Poultry | 140 | 0.92 (1.11) | 88 | 0.88 (1.13) | 0.04 [-0.18, 0.26] | 0.71 |
| Dogs/cats | 107 | 0.82 (1.11) | 121 | 0.97 (1.12) | -0.15 [-0.46, 0.16] | 0.35 |
| Animals observed within 2x2m of soil sampling area | 15 | 1.05 (1.13) | 213 | 0.89 (1.12) | 0.15 [-0.40, 0.71] | 0.59 |
| Animal feces observed in compound | 18 | 0.81 (1.08) | 210 | 0.91 (1.12) | -0.10 [-0.68, 0.48] | 0.73 |
| Animals enclosed during day | 7 | 0.66 (0.77) | 48 | 1.13 (1.18) | -0.46 [-1.19, 0.27] | 0.22 |
| Animals enclosed at night | 12 | 0.16 (0.55) | 43 | 1.32 (1.14) | **-1.15 [-1.64, -0.67]** | **<0.0005** |
| Household gave antibiotics in last 4 weeks to: |  |  |  |  |  |  |
| Animals | 16 | 0.83 (1.27) | 39 | 1.16 (1.08) | -0.34 [-1.01, 0.34] | 0.33 |
| Poultry | 8 | 0.87 (1.21) | 47 | 1.10 (1.14) | -0.23 [-1.15, 0.69] | 0.62 |
| Dogs/cats | 8 | 0.79 (1.40) | 47 | 1.11 (1.10) | -0.33 [-1.42, 0.76] | 0.56 |
| **Child health** |  |  |  |  |  |  |
| Any child in household had in last 7 days: |  |  |  |  |  |  |
| Diarrhea | 47 | 0.94 (1.17) | 181 | 0.89 (1.11) | 0.05 [-0.36, 0.46] | 0.82 |
| Acute respiratory infection | 115 | 0.85 (1.11) | 113 | 0.96 (1.13) | -0.11 [-0.46, 0.23] | 0.51 |
| Acute respiratory infection with fever | 62 | 0.72 (1.08) | 166 | 0.97 (1.12) | -0.26 [-0.65, 0.14] | 0.21 |
| Fever | 85 | 0.77 (1.14) | 143 | **0.98 (1.10)** | **-0.21 [-0.50, 0.07]** | **0.14** |
| Any child in household used antibiotics in last 4 weeks | 84 | 0.74 (1.11) | 144 | **1.00 (1.11)** | **-0.26 [-0.57, 0.05]** | **0.10** |
| **Environmental factors** |  |  |  |  |  |  |
| Soil sampling area in sunlight at time of collection | 88 | 0.72 (1.16) | 140 | 1.02 (1.07) | **-0.30 [-0.68, 0.08]** | **0.13** |
| Soil sampling area dry at time of collection | 51 | 0.21 (0.93) | 177 | 1.10 (1.09) | **-0.89 [-1.24, -0.54]** | **<0.0005** |
| Ambient temperature in top tertile | 57 | 0.69 (1.07) | 171 | 0.97 (1.12) | **-0.29 [-0.56, -0.01]** | **0.05** |
| Ambient humidity in top tertile | 109 | 0.98 (1.06) | 119 | 0.84 (1.16) | 0.14 [-0.27, 0.54] | 0.50 |

Log<sub>10</sub>-MPN: Log<sub>10</sub>transformed most-probable number cefotaxime-resistant *E. coli*. (Δlog<sub>10</sub>-MPN: difference between binary log<sub>10</sub> transformed most-probable number cefotaxime-resistant *E. coli*; SD: Standard Deviation; CI: Confidence Interval.

In multivariable models that adjusted for potential confounders, animal cohabitation intensity had a strong association with the abundance of cefotaxime-resistant *E. coli* in yard soil. Specifically, compared to households that did not own animals, households that owned animals and kept them enclosed at night had a significantly lower abundance of cefotaxime-resistant *E. coli* (Δlog_10_: -0.48 (-0.81, -0.15); p-value<0.005), whereas households that owned animals and did not enclose them at night had a higher abundance (Δlog_10_: 0.46 (0.20, 0.72); p-value<0.005) (Figure 2, S3 Table). Households also had a significantly lower abundance of cefotaxime-resistant *E. coli* if the soil was dry at the time of collection (Δlog_10_: -0.74 (-1.19, -0.29); p-value<0.005) and if the household was in the top asset-based household wealth quintile (Δlog_10_: - 0.72 (-1.13, -0.31); p-value<0.005) (Figure 2, S3 Table). In a separate multivariable model that considered poultry ownership rather than overall animal ownership, owning poultry was not associated with cefotaxime-resistant *E. coli* abundance, while the soil being dry and greater household wealth remained associated with lower cefotaxime-resistant *E. coli* abundance (S4 Table). Sanitation, aforementioned child health indicators, and ambient temperature did not have an association with cefotaxime-resistant *E. coli* abundance in yard soil in multivariable models (Figure 2, S3 Table, S4 Table).

**Figure 2.**
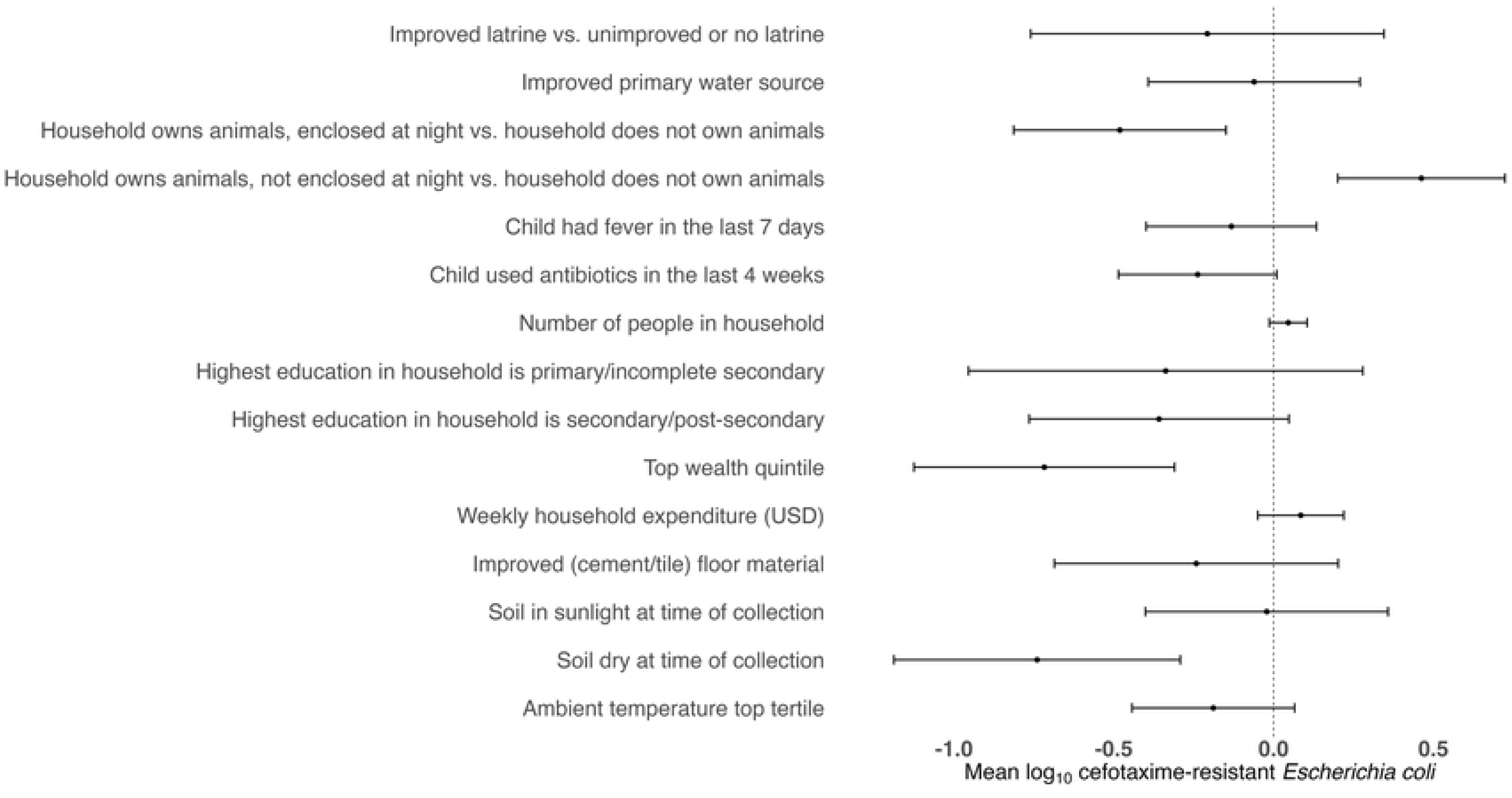
Forest plot of differences in mean log_10_-transformed most probable number (MPN) of cefotaxime-resistant *Escherichia coli* counts in yard soil associated with household-level indicators of sanitation, animal ownership and management, child health, and environmental variables in multivariable models. Models include variables that were associated with the outcome with a p-value of <0.20 in bivariate analyses and also control for improved primary water source, highest education in the household, asset- based household wealth quintile, weekly household expenditure, number of people in the household and improved floor material. Circles indicate point estimates and horizontal lines indicate 95% confidence intervals.

Findings for cefotaxime-resistant *E. coli* prevalence in yard soil were broadly similar to those for cefotaxime-resistant *E. coli* abundance. In bivariate models, households had significantly higher prevalence of cefotaxime-resistant *E. coli* in yard soil if they owned poultry (S5 Table). Additionally, while not significant, households appeared to have higher prevalence of cefotaxime-resistant *E. coli* if the household owned any animal, if children openly defecated, or if the ambient humidity was in the top tertile (S5 Table). In contrast, households had significantly lower prevalence of cefotaxime-resistant *E. coli* if animals were enclosed at night, if the soil was in sunlight at time of collection, if the soil was dry at time of collection, or if the ambient temperature was in the top tertile (S5 Table). While not statistically significant, households appeared to have lower prevalence of cefotaxime-resistant *E. coli* if they had a flush/pour flush latrine, the latrine was used by a single household, and any child in the household used antibiotics in the last four weeks (S5 Table). In multivariable models, any child in the household using antibiotics in the last four weeks (PR: 0.83 (0.70, 0.99); p-value=0.04), soil being dry at time of collection (PR: 0.38 (0.28, 0.51); p-value<0.0005), ambient humidity in the top tertile (PR: 0.83 (0.72, 0.96); p-value=0.01), and the household being in higher wealth quintiles (PR: 0.64 (0.46, 0.88); p-value=0.01) were associated with lower prevalence of cefotaxime-resistant *E. coli* in yard soil (S6 Table, S7 Table).

## Discussion

In our study in a peri-urban setting in Malawi, approximately two thirds of enrolled households with a child <5 years had cefotaxime-resistant *E. coli* in yard soil. Malawi is a low-income country and experiences high rates of AMR colonization both among humans and animals, consistent with prior evidence of widespread AMR in other low-income countries [62–65]. A previous study in southern Malawi detected ESBL-producing *E. coli* or *Klebsiella pneumoniae* in 30% of animal feces (n=290 samples) and 42% of human stools (n=1190 samples) [66]. Strengthening AMR surveillance and regulations, has been identified as a priority [67], and it is critical to investigate the local drivers of AMR transmission. In our study, unenclosed domestic animals were associated with higher abundance of cefotaxime-resistant *E. coli* in yard soil, while enclosed domestic animals, soil being dry and higher household wealth were associated with lower abundance, and household sanitation and antibiotic use were not associated with the prevalence or abundance of cefotaxime-resistant *E. coli* in yard soil.

Previous studies have identified soil as an important reservoir for AMR [21,28,29,68] and reported a range of prevalence values for antimicrobial-resistant *E. coli* in domestic, environmental and agricultural soils, including 14% of soil samples from a river basin in Tanzania [31], 42% of yard soil samples in Bangladesh [28], 68% of farm soil samples in Nigeria [30] and 89% of indoor soil floors in Bangladesh [27]. Specifically in Malawi, 77% of farm soil samples positive for *E. coli* showed cefotaxime resistance [69], and ESBL-producing bacteria (*E. coli, K. pneumoniae*) were found in 60% of soil samples from play and waste deposition areas in peri-urban Malawi [68]. Our finding of 68% prevalence in yard soil among peri-urban Malawian households is consistent with prior evidence. Recent studies in Malawi have also detected AMR in other environmental matrices such as farmland and household soil, vegetables, drains, standing water, and household drinking water [52,68,69]. However, in additional data from our study, the prevalence of cefotaxime-resistant *E. coli* in drinking water (8.4%) was substantially lower than what we detected in yard soil [52], pointing to soil as a notable reservoir. Children <5 years in our study were reported to play, eat, sleep and crawl on the ground outside, presenting opportunities for soil exposure. Given previous evidence on the contribution of direct and indirect soil ingestion to child exposure to fecal contamination [37], our findings suggest that domestic soils can be a source of AMR exposure for young children.

As environmental pathways are increasingly recognized to facilitate AMR transmission, understanding drivers of AROs in environmental matrices is critical. Here, animal cohabitation, specifically nighttime enclosure of domestic animals, had a strong association with cefotaxime-resistant *E. coli* abundance in yard soil. Domestic animals often share living space with household members in low-income countries [27,70,71]. Animal presence has previously been associated with soil contamination with fecal indicators and AROs [27,72],and living in close proximity to animals has been shown to increase the odds of being colonized with AROs [73]. Further, there is growing evidence on the contribution of specific animal management practices to soil contamination with AROs. In our study, unenclosed animals were associated with higher abundance of cefotaxime-resistant *E. coli* in yard soil while enclosed animals were associated lower abundance, compared to not owning animals. These findings align with a recent study in rural Bangladesh where households who kept their animals inside the home at night or let them roam free inside the home or in the compound had higher prevalence and abundance of cefotaxime-resistant *E. coli* on indoor soil floors [27]. This could be due to antibiotic use in backyard animal husbandry and resulting dissemination of AMR via fecal waste from unenclosed or freely roaming domestic animals. A study in Kenya found that 80% of survey respondents utilized antibiotics in domestic livestock practices [12], while 28% of households that owned an animal in our study gave antibiotics to their animals (primarily to chickens). A recent study examining antimicrobial usage polices in Malawi found that animal husbandry is an area where further policies are needed, as clinically-relevant antimicrobials are not regulated for animal usage, and animal feces disposal is unregulated [10]. However, we found no association between animal antibiotic use and cefotaxime-resistant *E. coli* in yard soil. We did not record the specific antibiotics used; it is possible that the antibiotics used for animals were not beta-lactams (e.g. tetracycline) and do not trigger ESBL production and/or cefotaxime resistance. Nonetheless, while antibiotic stewardship for domestic animals is recognized as an AMR control strategy [74], our findings highlight animal enclosure practices as an additional One Health approach to addressing AMR.

Poultry ownership was specifically associated with elevated cefotaxime-resistant *E. coli* in bivariate models in our study while this association was attenuated after controlling for potential confounders. Our findings are consistent with a 2017 study in Bangladesh that demonstrated domestic animals, especially chickens, contribute to fecal contamination in soil [32] and with a recent study that found associations between the number of chickens owned and cefotaxime-resistant *E. coli* on indoor soil floors [27]. In a study in Burkina Faso, children in households that owned poultry had greater odds of tetracycline resistance genes present in the gut compared to households that did not own poultry [75]. The specific contribution of poultry to fecal and AMR contamination in the domestic environment may be due to the difficulty in maintaining poultry enclosures [76]. Backyard poultry owners in low-income counties often prefer not to use enclosures because enclosed poultry require feed while free-range poultry can forage for food. Backyard poultry are also more likely to be allowed to roam freely inside living areas than larger animals, which are often tied up or kept in dedicated areas; roaming birds can contaminate the domestic environment as they scratch soil, forage and defecate. On the other hand, a study in Peru found that chicken corrals are vulnerable to contamination due to their structure, creating an intensive process for maintaining hygienic animal management infrastructure [77], and corralling chickens increased *Campylobacter* infections among young children, compared to letting them range free [78]. Our survey collected enclosure information broadly for all animals in the compound and did not record whether poultry specifically were confined in an enclosure. However, among households that used nighttime enclosures, 50% owned poultry while 42% owned poultry and at least one other type of animal. Analyzing enclosure types and practices that households use specifically for poultry may provide further insight on how poultry management may affect cefotaxime-resistant *E. coli* contamination in yard soil.

Vulnerable water and sanitation infrastructure has been identified as a key barrier in addressing AMR in Malawi, as weak infrastructure perpetuates fecal-oral transmission and may create an effective environment for AMR to disseminate [79]. In settings with inadequate sanitation, antibiotic residues, AROs, and ARGs can be disseminated into the environment human fecal waste [80]. In our study, cefotaxime-resistant *E. coli* abundance in soil was higher among households that had unimproved latrines shared by a higher number of users, and in unadjusted regression models, multiple household-level sanitation characteristics, such as having an improved or flush/pour flush latrine, were borderline associated with lower abundance of cefotaxime-resistant *E. coli* in yard soil, while open defecation by young children in the household was borderline associated with higher abundance. However, once we controlled for potential confounding factors such as primary water source and household wealth, these associations were attenuated. Similarly, existing evidence on the association between sanitation and AMR is mixed and vulnerable to confounding by wealth. In a global analysis using human gut metagenome data (i.e. genetic material within the human gut) from 26 countries in Africa, North America, Europe, South America, and Asia, access to sanitation infrastructure that effectively isolates waste from human contact was associated with lower abundance of ARGs in the gut metagenome [14]. A 2018 modelling study found that lacking WaSH infrastructure was associated with higher AMR colonization in humans [19]. A study in Malawi found that having inadequate sanitation facilities was associated with higher AMR levels in drains and soil [68]. However, much of the evidence on the association between sanitation status and AMR comes from observational studies, and it is possible that the findings are confounded by wealth (i.e. lower AMR in households/regions with better sanitation reflecting overall better health from higher wealth). In a randomized controlled trial in Bangladesh, on-site sanitation improvements did not decrease fecal contamination of courtyard soil [40], and there is mixed evidence on how effectively on-site sanitation systems isolate fecal waste from the environment in low-income countries [81]. Taken together with this prior evidence, our findings suggest that household-level sanitation practices may not fully capture drivers of cefotaxime resistance in environmental compartments such as soil; integrating other variables, such as community-level infrastructure and practices, environmental factors and animal husbandry, may identify critical additional drivers [82]. Additionally, future randomized sanitation trials that measure AMR in the environment or among residents as an outcome can help generate unconfounded evidence on the relationship between sanitation status and AMR.

In our analysis, child health variables such as diarrheal and respiratory infections, which are common drivers of pediatric antibiotic use, were not associated with cefotaxime-resistant *E. coli* in yard soil. In bivariate models, child antibiotic use in the last month was associated with lower cefotaxime-resistant *E. coli* prevalence and borderline associated with lower cefotaxime-resistant *E. coli* abundance in soil (p=0.06). However, in multivariable models, this relationship was attenuated. Notably, approximately a third of children in our study used antibiotics in the last month. The most common antibiotics used for children were amoxicillin and trimethoprim/ sulfamethoxazole. While trimethoprim/sulfamethoxazole belong to the sulfonamide class of antibiotics [83], amoxicillin belongs to the penicillin class and is a beta-lactam antibiotic like cefotaxime [84]. Therefore, frequent amoxicillin use by children would be expected to be associated with ESBL-production and/or cefotaxime resistance in children’s guts and consequently in domestic soils, given that 25% of children in our study open defecated within the compound. Our finding of no association between child antibiotic use and cefotaxime resistance in yard soil supports existing evidence that environmental transmission may contribute more to human colonization with AMR than the direct consumption of antimicrobials [19]. While we did not record antibiotic consumption by household members other than children <5 years, our findings suggest that factors outside of human consumption of antibiotics may be the dominant drivers of cefotaxime resistance in the home environment in peri-urban Malawi.

Some environmental factors such as soil being dry and in sunlight were associated with lower cefotaxime-resistant *E. coli* abundance in yard soil in our study. In contrast, while high ambient temperature was associated with lower cefotaxime-resistant *E. coli* abundance in bivariate models, this relationship was attenuated in multivariable models, and ambient humidity was not associated with cefotaxime-resistant *E. coli* abundance in bivariate or multivariate models. Our findings are consistent with previous evidence on the effect of moisture on *E. coli* and antimicrobial behavior. *E. coli* proliferates in soil under favorable environmental conditions, including higher moisture content [28,85]. Sunlight can inactivate microorganisms [86] and may break down antibiotic residues in soil; when exposed to sunlight, oxytetracycline and tetracycline, common broad-spectrum antibiotics used in humans and animals [87], lose antibiotic potential [88] and therefore may not trigger resistance. On the other hand, our findings differ from previous evidence on the relationship between ambient temperature/humidity and AMR. In a global study utilizing AMR databases, AMR colonization in inpatients was more common in areas with warmer temperatures [19]. An ecological study in China utilizing human clinical data found that, although ambient humidity confounds with ambient temperature, the median relative humidity of the area may influence the proliferation of AMR alongside ambient temperatures [60]. Our findings of no association with these weather parameters may reflect our data collection period during the cool and dry winter season in Malawi. In contrast, associations between ambient temperature and AMR in human clinical data have been derived from multi-year simulation studies, which better capture temperature variations [89]. We also did not analyze soil temperature specifically, which may influence the survival and proliferation of ARGs in soil [90]. Understanding how weather affects AROs is critical, given increasing temperatures and rainfall intensity in many world regions where AMR is common.

Our study had some limitations. First, we solely measured cefotaxime-resistant *E. coli*, which may not capture other AMR classes or specific resistant pathogens. Cefotaxime is a third generation cephalosporin and a broad-spectrum antibiotic [50,91]. Cefotaxime resistance is conferred by ESBL production; ESBL-producing organisms have multi-drug resistance against penicillins and cephalosporins, including cefotaxime [92]. Therefore, our study used a validated protocol [55] to quantify cefotaxime-resistant *E. coli* in yard soil as a proxy for ESBL-producing *E. coli* [50], which is widely monitored as a sentinel organism for AMR in environmental matrices, such as in the WHO’s Tricycle global AMR surveillance protocol [51]. We also did not measure cefotaxime-resistant *E. coli* or other AROs in human or animal stool to corroborate the associations with sanitation and animal management practices. Additionally, our cross-sectional data collection only captured the dry/cold season in Malawi; the abundance of cefotaxime-resistant *E. coli* in yard soil and factors associated with it may differ during warm/rainy seasons. Further, due to our observational cross-sectional study design, we could only establish association, not causation, between household and environmental variables and cefotaxime-resistant *E. coli* in yard soil. While we accounted for a range of potential confounders (water source, education, socio-demographics), the possibility of confounding remains. Finally, we collected self-reported data on household practices using a structured questionnaire. Respondents may remember events and answer survey questions inaccurately. However, we have no reason to expect that any such misreporting would be differential with respect to our study exposure and outcomes variables; any non-differential misreporting would be expected to attenuate the observed associations.

## Conclusion

Findings from this study suggest that while sanitation, child health characteristics and antibiotic use did not influence cefotaxime-resistant *E. coli* in yard soil, environmental and animal husbandry characteristics were significantly associated with the abundance and/or prevalence of cefotaxime-resistant *E. coli* in this peri-urban study setting. Results highlight the importance of taking a One Health approach to incorporate factors related to animals and the environment in global AMR mitigation strategies. Future studies should investigate the influence of specific animal husbandry practices, including animal cohabitation intensity and animal types, on cefotaxime-resistant *E. coli* in yard soil, other matrices in the domestic environment and among household members.

## Data Availability

Upon acceptance for publication, de-identified data will be publicly available without restriction at the following link: https://osf.io/5rg6e/overview?view_only=c912a26fa5a24a6f885b085f8edeaf8d

## Supporting Information Captions

**S1 Table. Socio-demographics and water, sanitation and hygiene conditions of enrolled households**

**S2 Table. Child health outcomes, antibiotic use and soil exposure**

**S1 Figure. Moisture content (%) of soil, stratified by enumerator observation if the soil was wet/dry at time of collection.** The X-axis reflects whether or not the soil was dry an=t time of collection, and the Y-axis reflects the average moisture content (%) of the sample. The bottommost and topmost horizontal lines represent the lower and upper limits (excluding outliers), the bottom and top borders of the box represent the first and third quartiles, and the bold horizontal line represents the median of moisture content (%).

**S2 Figure. Mean log_10_-transformed most probable number (MPN) of cefotaxime-resistant *Escherichia coli* (CR-EC) among cross-categories of animal ownership and husbandry characteristics.** Y-axis variables include animals the household or compound owns, including any animals, dogs/cats, and poultry. X-axis variables include specific animal husbandry characteristics, including if the animal is enclosed in the daytime/nighttime, household administers antibiotics to animals, and animal feces were observed in the compound during sampling. The heatmap gradient reflects the mean log10-transformed MPN of CR-EC in yard soil, where darker colors represent higher MPN values. Boundaries indicate complementary variables.

**S3 Table. Adjusted associations between household environmental characteristics, animal ownership and abundance of cefotaxime-resistant *E. coli* in yard soil.** Models include variables that were associated with the outcome with a p-value of <0.20 in bivariate analyses and also control for household’s primary water source, education, socioeconomics, household size, and indoor floor material. Bolded values indicate associations with p-value <0.05.

**S4 Table. Adjusted associations between household environmental characteristics, poultry ownership and abundance of cefotaxime-resistant *E. coli* in yard soil.** Models include variables that were associated with the outcome with a p-value of <0.20 in bivariate analyses and also control for household’s primary water source, education, socioeconomics, household size, and indoor floor material. Bolded values indicate associations with p-value <0.05.

**S5 Table. Bivariate associations between household environmental characteristics and prevalence of cefotaxime-resistant *E. coli* in yard soil.** Bolded values indicate associations with p-value <0.05.

**S6 Table. Adjusted associations between household environmental characteristics, animal ownership and prevalence of cefotaxime-resistant *E. coli* in yard soil.** Models include variables that were associated with the outcome with a p-value of <0.20 in bivariate analyses and also control for household’s primary water source, education, socioeconomics, household size, and indoor floor material. Bolded values indicate associations with p-value <0.05.

**S7 Table. Adjusted associations between household environmental characteristics, poultry ownership and prevalence of cefotaxime-resistant *E. coli* in yard soil.** Models include variables that were associated with the outcome with a p-value of <0.20 in bivariate analyses and also control for household’s primary water source, education, socioeconomics, household size, and indoor floor material. Bolded values indicate associations with p-value <0.05.

## Notes

### Competing Interest Statement

The authors have declared no competing interest.

### Funding Statement

Grants that supported this work include the following: National Science Foundation 1951006 (Workman), National Science Foundation 1950212 (Harris), National Science Foundation 2236372 (de los Reyes III), and United States Department of Education GAANN P200A240029. The funders had no role in study design, data collection and analysis, decision to publish, or preparation of the manuscript.

### Author Declarations

We obtained IRB approval from the Research Ethics Committee (MZUNIREC) of Mzuzu University (MZUNIREC/DOR/24/38) and the Ethics committee/IRB of University of North Carolina Greensboro (UNC-Greensboro IRB #: 20-0245).

## References

1. Murray CJL, Ikuta KS, Sharara F, Swetschinski L, Robles Aguilar G, Gray A, et al. Global burden of bacterial antimicrobial resistance in 2019: a systematic analysis. The Lancet. 2022;399: 629–655. doi:10.1016/S0140-6736(21)02724-0

2. O’Neill J. Tackling Drug-Resistant Infections Globally: Final Report and Recommendations. 2016 May. Available: https://amr-review.org/sites/default/files/160525_Final%20paper_with%20cover.pdf

3. Naghavi M, Vollset SE, Ikuta KS, Swetschinski LR, Gray AP, Wool EE, et al. Global burden of bacterial antimicrobial resistance 1990–2021: a systematic analysis with forecasts to 2050. The Lancet. 2024;404: 1199–1226. doi:10.1016/S0140-6736(24)01867-1

4. Founou RC, Founou LL, Essack SY. Clinical and economic impact of antibiotic resistance in developing countries: A systematic review and meta-analysis. Butaye P, editor. PLOS ONE. 2017;12: e0189621. doi:10.1371/journal.pone.0189621

5. Koukoubani T, Makris D, Daniil Z, Paraforou T, Tsolaki V, Zakynthinos E, et al. The role of antimicrobial resistance on long-term mortality and quality of life in critically ill patients: a prospective longitudinal 2-year study. Health Qual Life Outcomes. 2021;19: 72. doi:10.1186/s12955-021-01712-0

6. Matos ECOD, Andriolo RB, Rodrigues YC, Lima PDLD, Carneiro ICDRS, Lima KVB. Mortality in patients with multidrug-resistant Pseudomonas aeruginosa infections: a meta-analysis. Rev Soc Bras Med Trop. 2018;51: 415–420. doi:10.1590/0037-8682-0506-2017

7. Shrestha P, Cooper BS, Coast J, Oppong R, Do Thi Thuy N, Phodha T, et al. Enumerating the economic cost of antimicrobial resistance per antibiotic consumed to inform the evaluation of interventions affecting their use. Antimicrob Resist Infect Control. 2018;7. doi:10.1186/s13756-018-0384-3

8. Tompkins K, Juliano JJ, van Duin D. Antimicrobial Resistance in Enterobacterales and Its Contribution to Sepsis in Sub-saharan Africa. Front Med. 2021;8: 615649. doi:10.3389/fmed.2021.615649

9. Edwards T, Williams CT, Olwala M, Andang’o P, Otieno W, Nalwa GN, et al. Molecular surveillance reveals widespread colonisation by carbapenemase and extended spectrum beta-lactamase producing organisms in neonatal units in Kenya and Nigeria. Antimicrob Resist Infect Control. 2023;12: 14. doi:10.1186/s13756-023-01216-0

10. Mhone AL, Muloi DM, Moodley A. One Health: governance and regulatory framework for antimicrobial use in Malawi. Sci One Health. 2025;4: 100119. doi:10.1016/j.soh.2025.100119

11. Limwado GD, Aron MB, Mpinga K, Phiri H, Chibvunde S, Banda C, et al. Prevalence of antibiotic self-medication and knowledge of antimicrobial resistance among community members in Neno District rural Malawi: A cross-sectional study. IJID Reg. 2024;13: 100444. doi:10.1016/j.ijregi.2024.100444

12. Rware H, Monica KK, Idah M, Fernadis M, Davis I, Buke W, et al. Examining antibiotic use in Kenya: farmers’ knowledge and practices in addressing antibiotic resistance. CABI Agric Biosci. 2024 [cited 23 July 2025]. doi:10.1186/s43170-024-00223-4

13. World Health Organization. Antimicrobial Resistance. World Health Organization; 2023. Available: https://www.who.int/news-room/fact-sheets/detail/antimicrobial-resistance

14. Fuhrmeister ER, Harvey AP, Nadimpalli ML, Gallandat K, Ambelu A, Arnold BF, et al. Evaluating the relationship between community water and sanitation access and the global burden of antibiotic resistance: an ecological study. Lancet Microbe. 2023;4: e591–e600. doi:10.1016/S2666-5247(23)00137-4

15. Zhou Z, Chen H. Evaluating human exposure to antibiotic resistance genes. Biosaf Health. 2024;6: 98–100. doi:10.1016/j.bsheal.2024.02.005

16. Centers for Disease Control and Prevention. The Environment and Food Production – Causes of Antimicrobial Resistance. Centers for Disease Control and Prevention; 2024. Available: https://www.cdc.gov/antimicrobial-resistance/causes/environmental-food.html

17. Lienert J, Güdel K, Escher BI. Screening Method for Ecotoxicological Hazard Assessment of 42 Pharmaceuticals Considering Human Metabolism and Excretory Routes. Environ Sci Technol. 2007;41: 4471–4478. doi:10.1021/es0627693

18. Peng S, Zhang H, Song D, Chen H, Lin X, Wang Y, et al. Distribution of antibiotic, heavy metals and antibiotic resistance genes in livestock and poultry feces from different scale of farms in Ningxia, China. J Hazard Mater. 2022;440: 129719. doi:10.1016/j.jhazmat.2022.129719

19. Collignon P, Beggs JJ, Walsh TR, Gandra S, Laxminarayan R. Anthropological and socioeconomic factors contributing to global antimicrobial resistance: a univariate and multivariable analysis. Lancet Planet Health. 2018;2: e398–e405. doi:10.1016/S2542-5196(18)30186-4

20. Blair JMA, Webber MA, Baylay AJ, Ogbolu DO, Piddock LJV. Molecular mechanisms of antibiotic resistance. Nat Rev Microbiol. 2015;13: 42–51. doi:10.1038/nrmicro3380

21. Zhu Y-G, Zhao Y, Zhu D, Gillings M, Penuelas J, Ok YS, et al. Soil biota, antimicrobial resistance and planetary health. Environ Int. 2019;131: 105059. doi:10.1016/j.envint.2019.105059

22. Delgado-Baquerizo M, Hu H-W, Maestre FT, Guerra CA, Eisenhauer N, Eldridge DJ, et al. The global distribution and environmental drivers of the soil antibiotic resistome. Microbiome. 2022;10. doi:10.1186/s40168-022-01405-w

23. Imran Md, Das KR, Naik MM. Co-selection of multi-antibiotic resistance in bacterial pathogens in metal and microplastic contaminated environments: An emerging health threat. Chemosphere. 2019;215: 846–857. doi:10.1016/j.chemosphere.2018.10.114

24. Pagaling E, Hough R, Avery L, Robinson L, Freitag T, Coull M, et al. Antibiotic resistance patterns in soils across the Scottish landscape. Commun Earth Environ. 2023;4: 403. doi:10.1038/s43247-023-01057-0

25. Forsberg KJ, Reyes A, Wang B, Selleck EM, Sommer MOA, Dantas G. The shared antibiotic resistome of soil bacteria and human pathogens. Science. 2012;337: 1107–1111. doi:10.1126/science.1220761

26. Nguyen AT, Ratnasiri K, Barratt Heitmann G, Tazin S, Anderson C, Hanif S, et al. Potential pathogens and antimicrobial resistance genes in household environments: a study of soil floors and cow dung in rural Bangladesh. Dozois CM, editor. Appl Environ Microbiol. 2025;91: e00669–25. doi:10.1128/aem.00669-25

27. Ercumen A, Hossain MdS, Tabassum T, Haque A, Rahman A, Rahman MdH, et al. Dirt floors and domestic animals are associated with soilborne exposure to antimicrobial resistant E. coli in rural Bangladeshi households. Environ Sci Technol. 2025;59. 10.1021/acs.est.5c10329

28. Montealegre MC, Roy S, Böni F, Hossain MI, Navab-Daneshmand T, Caduff L, et al. Risk Factors for Detection, Survival, and Growth of Antibiotic-Resistant and Pathogenic Escherichia coli in Household Soils in Rural Bangladesh. Vieille C, editor. Appl Environ Microbiol. 2018;84: e01978–18. doi:10.1128/AEM.01978-18

29. Hartinger SM, Medina-Pizzali ML, Salmon-Mulanovich G, Larson AJ, Pinedo-Bardales M, Verastegui H, et al. Antimicrobial Resistance in Humans, Animals, Water and Household Environs in Rural Andean Peru: Exploring Dissemination Pathways through the One Health Lens. Int J Environ Res Public Health. 2021;18: 4604. doi:10.3390/ijerph18094604

30. Igbinosa EO, Beshiru A, Igbinosa IH, Cho G-S, Franz CMAP. Multidrug-resistant extended spectrum β-lactamase (ESBL)-producing Escherichia coli from farm produce and agricultural environments in Edo State, Nigeria. Aworh MK, editor. PLOS ONE. 2023;18: e0282835. doi:10.1371/journal.pone.0282835

31. Kimera ZI, Mgaya FX, Mshana SE, Karimuribo ED, Matee MIN. Occurrence of Extended Spectrum Beta Lactamase (ESBL) Producers, Quinolone and Carbapenem Resistant Enterobacteriaceae Isolated from Environmental Samples along Msimbazi River Basin Ecosystem in Tanzania. Int J Environ Res Public Health. 2021;18: 8264. doi:10.3390/ijerph18168264

32. Ercumen A, Pickering AJ, Kwong LH, Arnold BF, Parvez SM, Alam M, et al. Animal Feces Contribute to Domestic Fecal Contamination: Evidence from E. coli Measured in Water, Hands, Food, Flies, and Soil in Bangladesh. Environ Sci Technol. 2017;51: 8725–8734. doi:10.1021/acs.est.7b01710

33. Wang Y, Moe CL, Null C, Raj SJ, Baker KK, Robb KA, et al. Multipathway Quantitative Assessment of Exposure to Fecal Contamination for Young Children in Low-Income Urban Environments in Accra, Ghana: The SaniPath Analytical Approach. Am J Trop Med Hyg. 2017;97: 1009–1019. doi:10.4269/ajtmh.16-0408

34. Bauza V, Ocharo RM, Nguyen TH, Guest JS. Soil Ingestion is Associated with Child Diarrhea in an Urban Slum of Nairobi, Kenya. Am J Trop Med Hyg. 2017;96: 569–575. doi:10.4269/ajtmh.16-0543

35. Shivoga WA, Moturi WN. Geophagia as a risk factor for diarrhoea. J Infect Dev Ctries. 2009;3: 094–098. doi:10.3855/jidc.55

36. Kwong, Ercumen A, Pickering A, Unicomb L, Davis J, Luby S. Hand- and Object-Mouthing of Rural Bangladeshi Children 3–18 Months Old. Int J Environ Res Public Health. 2016;13: 563. doi:10.3390/ijerph13060563

37. Kwong, Ercumen A, Pickering AJ, Unicomb L, Davis J, Leckie JO, et al. Soil ingestion among young children in rural Bangladesh. J Expo Sci Environ Epidemiol. 2021;31: 82–93. doi:10.1038/s41370-019-0177-7

38. Sammarro M, Rowlingson B, Cocker D, Chidziwisano K, Jacob ST, Kajumbula H, et al. Risk Factors, Temporal Dependence, and Seasonality of Human Extended-Spectrum β-Lactamases-Producing Escherichia coli and Klebsiella pneumoniae Colonization in Malawi: A Longitudinal Model-Based Approach. Clin Infect Dis. 2023;77: 1–8. 10.1093/cid/ciad117

39. Pickering AJ, Julian TR, Marks SJ, Mattioli MC, Boehm AB, Schwab KJ, et al. Fecal Contamination and Diarrheal Pathogens on Surfaces and in Soils among Tanzanian Households with and without Improved Sanitation. Environ Sci Technol. 2012;46: 5736–5743. doi:10.1021/es300022c

40. Ercumen A, Pickering AJ, Kwong LH, Mertens A, Arnold BF, Benjamin-Chung J, et al. Do Sanitation Improvements Reduce Fecal Contamination of Water, Hands, Food, Soil, and Flies? Evidence from a Cluster-Randomized Controlled Trial in Rural Bangladesh. Environ Sci Technol. 2018;52: 12089–12097. doi:10.1021/acs.est.8b02988

41. World Health Organization. Technical brief on water, sanitation, hygiene (WASH) and wastewater management to prevent infections and reduce the spread of antimicrobial resistance (AMR). World Health Organization; 2020 Nov. Available: https://www.who.int/publications/i/item/9789240006416

42. Peng J-J, Balasubramanian B, Ming Y-Y, Niu J-L, Yi C-M, Ma Y, et al. Identification of antimicrobial resistance genes and drug resistance analysis of Escherichia coli in the animal farm environment. J Infect Public Health. 2021;14: 1788–1795. doi:10.1016/j.jiph.2021.10.025

43. Scicchitano D, Leuzzi D, Babbi G, Palladino G, Turroni S, Laczny CC, et al. Dispersion of antimicrobial resistant bacteria in pig farms and in the surrounding environment. Anim Microbiome. 2024;6: 17. doi:10.1186/s42523-024-00305-8

44. Shen C, He M, Zhang J, Liu J, Su J, Dai J. Effects of the coexistence of antibiotics and heavy metals on the fate of antibiotic resistance genes in chicken manure and surrounding soils. Ecotoxicol Environ Saf. 2023;263: 115367. doi:10.1016/j.ecoenv.2023.115367

45. Shen D, Li C, Guo Z. Dynamics of antibiotic resistance in poultry farms via multivector analysis. Poult Sci. 2025;104: 104673. doi:10.1016/j.psj.2024.104673

46. Zhou B, Wang C, Zhao Q, Wang Y, Huo M, Wang J, et al. Prevalence and dissemination of antibiotic resistance genes and coselection of heavy metals in Chinese dairy farms. J Hazard Mater. 2016;320: 10–17. doi:10.1016/j.jhazmat.2016.08.007

47. Swarthout JM, Chan EMG, Garcia D, Nadimpalli ML, Pickering AJ. Human Colonization with Antibiotic-Resistant Bacteria from Nonoccupational Exposure to Domesticated Animals in Low- and Middle-Income Countries: A Critical Review. Environ Sci Technol. 2022;56: 14875–14890. doi:10.1021/acs.est.2c01494

48. Shawver S, Ishii S, Strickland MS, Badgley B. Soil type and moisture content alter soil microbial responses to manure from cattle administered antibiotics. Environ Sci Pollut Res Int. 2024;31: 27259–27272. doi:10.1007/s11356-024-32903-z

49. Ishii S, Ksoll WB, Hicks RE, Sadowsky MJ. Presence and growth of naturalized Escherichia coli in temperate soils from Lake Superior watersheds. Appl Environ Microbiol. 2006;72: 612–621. doi:10.1128/AEM.72.1.612-621.2006

50. Carmine AA, Brogden RN, Heel RC, Speight TM, Avery GS. Cefotaxime A review of its Antibacterial Activity, Pharmacological Properties and Therapeutic Use: Drugs. 1983;25: 223–289. doi:10.2165/00003495-198325030-00001

51. World Health Organization. WHO integrated global surveillance on ESBL-producing E. coli using a “One Health” approach: implementation and opportunities. World Health Organization; 2021 Mar. Available: https://www.who.int/publications/i/item/9789240021402

52. Niven CG, Clark B, Floess E, Chirwa B, Matekenya M, Budden E, et al. Associations between water supply intermittencies and drinking water quality, child health, and caregiver emotional stress in peri-urban Malawi. 2026. doi:10.1021/acs.est.5c08820

53. JMP. Core questions on drinking water, sanitation and hygiene for household surveys: 2018 update. United Nations Children’s Fund (UNICEF) and World Health Organization; 2018.

54. Blantyre Weather History. Weather API; Available: https://www.weatherapi.com/history/q/blantyre-1598090?loc=1598090

55. Hornsby G, Ibitoye TD, Keelara S, Harris A. Validation of a modified IDEXX defined-substrate assay for detection of antimicrobial resistant *E. coli* in environmental reservoirs. Environ Sci Process Impacts. 2023;25: 37–43. doi:10.1039/D2EM00189F

56. Weldelberger J, Campbell K. Non-detect data in environmental investigations. Toronto, Canada: Los Alamos National Laboratory; 1994. Available: https://www.osti.gov/servlets/purl/10156972

57. World Health Organization (WHO), United Nations Children’s Fund (UNICEF). Progress on household drinking water, sanitation and hygiene 2000–2024: special focus on inequalities. Geneva; 2025 Aug.

58. Ashraf S, Islam M, Unicomb L, Rahman M, Winch PJ, Arnold BF, et al. Effect of Improved Water Quality, Sanitation, Hygiene and Nutrition Interventions on Respiratory Illness in Young Children in Rural Bangladesh: A Multi-Arm Cluster-Randomized Controlled Trial. Am J Trop Med Hyg. 2020;102: 1124–1130. doi:10.4269/ajtmh.19-0769

59. Martel P. Review of options for reporting water, sanitation and hygiene coverage by wealth quintile. U N Child Fund UNICEF Data Anal Sect Div Data Res Policy UNICEF N Y. 2016;No. 4. doi:https://mics.unicef.org/sites/mics/files/2024-05/MICS%20Methodological%20Paper%204.pdf

60. Li W, Liu C, Ho HC, Shi L, Zeng Y, Yang X, et al. Estimating the effect of increasing ambient temperature on antimicrobial resistance in China: A nationwide ecological study with the difference-in-differences approach. Sci Total Environ. 2023;882: 163518. doi:10.1016/j.scitotenv.2023.163518

61. Muloi DM, Hassell JM, Wee BA, Ward MJ, Bettridge JM, Kivali V, et al. Genomic epidemiology of Escherichia coli: antimicrobial resistance through a One Health lens in sympatric humans, livestock and peri-domestic wildlife in Nairobi, Kenya. BMC Med. 2022;20: 471. doi:10.1186/s12916-022-02677-7

62. Bizimungu O, Crook P, Babane JF, Bitunguhari L. The prevalence and clinical context of antimicrobial resistance amongst medical inpatients at a referral hospital in Rwanda: a cohort study. Antimicrob Resist Infect Control. 2024;13: 22. doi:10.1186/s13756-024-01384-7

63. Cooper LN, Beauchamp AM, Ingle TA, Diaz MI, Wakene AD, Katterpalli C, et al. Socioeconomic Disparities and the Prevalence of Antimicrobial Resistance. Clin Infect Dis. 2024;79: 1346–1353. doi:10.1093/cid/ciae313

64. Jajoo M, Manchanda V, Chaurasia S, Sankar MJ, Gautam H, Agarwal R, et al. Alarming rates of antimicrobial resistance and fungal sepsis in outborn neonates in North India. PloS One. 2018;13: e0180705. doi:10.1371/journal.pone.0180705

65. Johnson J, Robinson ML, Rajput UC, Valvi C, Kinikar A, Parikh TB, et al. High Burden of Bloodstream Infections Associated With Antimicrobial Resistance and Mortality in the Neonatal Intensive Care Unit in Pune, India. Clin Infect Dis Off Publ Infect Dis Soc Am. 2021;73: 271–280. doi:10.1093/cid/ciaa554

66. Cocker D, Chidziwisano K, Mphasa M, Mwapasa T, Lewis JM, Rowlingson B, et al. Investigating One Health risks for human colonisation with extended spectrum β-lactamase-producing Escherichia coli and Klebsiella pneumoniae in Malawian households: a longitudinal cohort study. Lancet Microbe. 2023;4: e534–e543. doi:10.1016/S2666-5247(23)00062-9

67. World Health Organization. Malawi national action plan on antimicrobial resistance: review of progress in the human health sector. Geneva: World Health Organization; 2022.

68. Mwapasa T, Chidziwisano K, Mphasa M, Cocker D, Rimella L, Amos S, et al. Key environmental exposure pathways to antimicrobial resistant bacteria in southern Malawi: A SaniPath approach. Sci Total Environ. 2024;945: 174142. doi:10.1016/j.scitotenv.2024.174142

69. Abraham A, Mtewa AG, Chiutula C, Mvula RLS, Maluwa A, Eregno FE, et al. Prevalence of Antibiotic Resistance Bacteria in Manure, Soil, and Vegetables in Urban Blantyre, Malawi, from a Farm-to-Fork Perspective. Int J Environ Res Public Health. 2025;22: 1273. doi:10.3390/ijerph22081273

70. Barnes AN, Mumma J, Cumming O. Role, ownership and presence of domestic animals in peri-urban households of Kisumu, Kenya. Zoonoses Public Health. 2018;65: 202–214. doi:10.1111/zph.12429

71. Passarelli S, Ambikapathi R, Gunaratna NS, Madzorera I, Canavan CR, Noor RA, et al. The role of chicken management practices in children’s exposure to environmental contamination: a mixed-methods analysis. BMC Public Health. 2021;21: 1097. doi:10.1186/s12889-021-11025-y

72. Ercumen A, Pickering AJ, Kwong LH, Arnold BF, Parvez SM, Alam M, et al. Animal Feces Contribute to Domestic Fecal Contamination: Evidence from *E. coli* Measured in Water, Hands, Food, Flies, and Soil in Bangladesh. Environ Sci Technol. 2017;51: 8725–8734. doi:10.1021/acs.est.7b01710

73. Kurowski KM, Marusinec R, Amato HK, Saraiva-Garcia C, Loayza F, Salinas L, et al. Social and Environmental Determinants of Community-Acquired Antimicrobial-Resistant Escherichia coli in Children Living in Semirural Communities of Quito, Ecuador. Am J Trop Med Hyg. 2021;105: 600–610. doi:10.4269/ajtmh.20-0532

74. Van Boeckel TP, Pires J, Silvester R, Zhao C, Song J, Criscuolo NG, et al. Global trends in antimicrobial resistance in animals in low- and middle-income countries. Science. 2019;365: eaaw1944. doi:10.1126/science.aaw1944

75. Brogdon JM, Sié A, Dah C, Ouermi L, Coulibaly B, Lebas E, et al. Poultry Ownership and Genetic Antibiotic Resistance Determinants in the Gut of Preschool Children. Am J Trop Med Hyg. 2021;104: 1768–1770. doi:10.4269/ajtmh.20-1384

76. Thomas ED, Kwong LH, Rimi NA, Shanta IS, Uddin MR, Khan S, et al. Who has sheds? Exploring practices and determinants of overnight housing for backyard poultry in rural Bangladesh to inform an intervention to limit exposure to poultry and poultry feces. Robinson J, editor. PLOS Glob Public Health. 2025;5: e0004929. doi:10.1371/journal.pgph.0004929

77. Harvey SA, Winch PJ, Leontsini E, Torres Gayoso C, López Romero S, Gilman RH, et al. Domestic poultry-raising practices in a Peruvian shantytown: implications for control of Campylobacter jejuni-associated diarrhea. Acta Trop. 2003;86: 41–54. doi:10.1016/S0001-706X(03)00006-8

78. Oberhelman RA, Gilman RH, Sheen P, Cordova J, Zimic M, Cabrera L, et al. AN INTERVENTION-CONTROL STUDY OF CORRALLING OF FREE-RANGING CHICKENS TO CONTROL CAMPYLOBACTER INFECTIONS AMONG CHILDREN IN A PERUVIAN PERIURBAN SHANTYTOWN. Am J Trop Med Hyg. 2006;74: 1054–1059. doi:10.4269/ajtmh.2006.74.1054

79. Kalitsilo L, Morse T, Feasy N, Malenga T, Kamthunzi V, Chidziwosano K. Water, sanitation and hygiene (WASH): a critical barrier against antimicrobial resistance (AMR) in Malawi. 2021 May.

80. Berglund B. Environmental dissemination of antibiotic resistance genes and correlation to anthropogenic contamination with antibiotics. Infect Ecol Epidemiol. 2015;5: 28564. doi:10.3402/iee.v5.28564

81. Sclar GD, Garn JV, Penakalapati G, Alexander KT, Krauss J, Freeman MC, et al. Effects of sanitation on cognitive development and school absence: A systematic review. Int J Hyg Environ Health. 2017;220: 917–927. doi:10.1016/j.ijheh.2017.06.010

82. Contreras JD, Islam M, Mertens A, Pickering AJ, Kwong LH, Arnold BF, et al. Influence of community-level sanitation coverage and population density on environmental fecal contamination and child health in a longitudinal cohort in rural Bangladesh. Int J Hyg Environ Health. 2022;245: 114031. doi:10.1016/j.ijheh.2022.114031

83. Hitchings GH. Mechanism of Action of Trimethoprim-Sulfamethoxazole--I. J Infect Dis. 1973;128: S433–S436. doi:10.1093/infdis/128.Supplement_3.S433

84. Akhavan BJ, Khanna NR, Vijhani P. Amoxicillin. StatPearls. Treasure Island (FL): StatPearls Publishing; 2025. Available: http://www.ncbi.nlm.nih.gov/books/NBK482250/

85. Ishii S, Yan T, Vu H, Hansen DL, Hicks RE, Sadowsky MJ. Factors Controlling Long-Term Survival and Growth of Naturalized Escherichia coli Populations in Temperate Field Soils. Microbes Environ. 2010;25: 8–14. doi:10.1264/jsme2.ME09172

86. Fahimipour AK, Hartmann EM, Siemens A, Kline J, Levin DA, Wilson H, et al. Daylight exposure modulates bacterial communities associated with household dust. Microbiome. 2018;6: 175. doi:10.1186/s40168-018-0559-4

87. Chopra I, Roberts M. Tetracycline antibiotics: mode of action, applications, molecular biology, and epidemiology of bacterial resistance. Microbiol Mol Biol Rev MMBR. 2001;65: 232–260 ; second page, table of contents. doi:10.1128/MMBR.65.2.232-260.2001

88. Khan SJ, Osborn AM, Eswara PJ. Effect of Sunlight on the Efficacy of Commercial Antibiotics Used in Agriculture. Front Microbiol. 2021;12: 645175. doi:10.3389/fmicb.2021.645175

89. Kou R, Li X, Wang H, Wu Y, Zou D, Hu J, et al. Panel data analysis of the effect of ambient temperature on antimicrobial resistance in China. Sci Rep. 2025;15: 18391. doi:10.1038/s41598-025-02861-8

90. Lin D, Du S, Zhao Z, Zhang T, Wang L, Zhang Q, et al. Climate warming fuels the global antibiotic resistome by altering soil bacterial traits. Nat Ecol Evol. 2025 [cited 23 July 2025]. doi:10.1038/s41559-025-02740-5

91. Padda IS, Nagalli S. Cefotaxime. StatPearls. Treasure Island (FL): StatPearls Publishing; 2025. Available: http://www.ncbi.nlm.nih.gov/books/NBK560653/

92. Shaikh S, Fatima J, Shakil S, Rizvi SMD, Kamal MA. Antibiotic resistance and extended spectrum beta-lactamases: Types, epidemiology and treatment. Saudi J Biol Sci. 2015;22: 90–101. doi:10.1016/j.sjbs.2014.08.002

